# PainWaive: A Consumer-grade Digitally Delivered EEG Neurofeedback Intervention for Chronic Low Back Pain

**DOI:** 10.64898/2026.03.26.26349247

**Authors:** Negin Hesam-Shariati, Ekaterina Ermolenko, Nahian S. Chowdhury, Pauline Zahara, Kevin Yi Chen, Chin-Teng Lin, Toby Newton-John, Sylvia M. Gustin

**Affiliations:** NeuroRecovery Research Hub, School of Psychology, University of New South Wales, Sydney, NSW, AU; Centre for Pain IMPACT, Neuroscience Research Australia, Sydney, NSW, AU; Human-centric Artificial Intelligence Centre, Australian AI Institute, FEIT, University of Technology Sydney, Sydney, Australia; Graduate School of Health, University of Technology Sydney, Sydney, NSW, AU

## Abstract

Chronic low back pain (CLBP) is persistent and refractory, affecting 20-30% of population worldwide. Neurofeedback has been explored as a potential non-pharmacological intervention for chronic pain, although evidence in CLBP remains limited. This study evaluated PainWaive, a consumer-grade digitally-delivered neurofeedback intervention targeting multiple pain-related frequency bands recorded over the sensorimotor cortex in individuals with CLBP.

In a multiple-baseline experimental design, four participants completed daily assessments of pain severity and pain interference during randomly-assigned baseline phases of 7, 10, 14, and 20 days, followed by 20 sessions of the PainWaive intervention over four weeks. Daily pain assessments continued during the post-intervention and follow-up phases. Participants rated PainWaive’s usability and acceptability at post-intervention. Anxiety, depression, wellbeing, and sleep disturbance were assessed at three timepoints.

Aggregated *Tau-U* analyses indicated a large effect (−0.67) on pain severity from baseline to intervention and very large from baseline to post-intervention (−0.92) and follow-up (−0.92) phases. Large effects (−0.63, −0.62, and −0.70) were also observed for pain interference. Individual-level analyses showed significant reductions across all participants, with visual inspection confirming progressive decreases over time. The intervention was rated usable and acceptable by all participants, while psychological outcomes were mixed and varied across participants.

The findings provide promising evidence that the PainWaive neurofeedback intervention may reduce pain severity and pain interference in some individuals with CLBP. By prioritising accessibility, usability, and self-administration, PainWaive supports a foundation for more patient-centred, technology-enabled approaches to chronic pain management. Further evaluation of this approach in randomised trials is required to establish efficacy.

## INTRODUCTION

Recent estimates from the Global Burden of Disease (GBD) study indicate that more than 600 million people were living with low back pain in 2020^1^, maintaining its position as the leading cause of global disability, with projections exceeding 800 million cases by 2050^1^. Evidence suggests that approximately one-third of people with low back pain continue to experience persistent pain and disability three months after onset^2^. Population studies indicate that chronic low back pain (CLBP) affects approximately 20% of adults aged 20–59 years^3^, with prevalence increasing with age to around 25–30% of individuals aged 60 years and older^3,4^. CLBP is often multifactorial, with biological, psychological, and social contributors. A substantial individuals with CLBP receive limited benefit from existing treatments, with pain remaining persistent and refractory^5^. This highlights the need to identify more effective treatment options.

Electroencephalographic (EEG) neurofeedback has been explored as a potential pain management intervention over the past few decades across a variety of chronic pain conditions, including CLBP^6–8^. Recent reviews^9–11^ suggest that EEG neurofeedback may offer therapeutic benefit for chronic pain; however, evidence specific to CLBP remains limited.

The rationale for using EEG neurofeedback in chronic pain is grounded in evidence that chronic pain reflects not only altered sensory processing but also maladaptive changes in peripheral and central nervous system pathways^12^. EEG provides a non-invasive method for detecting such alterations. Neurofeedback protocols used in CLBP and several other chronic pain studies have primarily targeted the alpha band (8–12 Hz)^6,8^ or infraslow frequencies (<0.1 Hz)^7^. However, emerging evidence indicates that chronic pain is characterised by alterations across multiple frequency bands, including reduced alpha power^12,13^ and increased theta and high-beta power^13^. In addition to frequency-specific alterations, a systematic review has demonstrated altered corticospinal and intracortical excitability in chronic pain populations, with consistent evidence of motor cortex disinhibition suggestive of disrupted GABA-mediated intracortical inhibition^14^. Given the central role of the sensorimotor cortex in integrating somatosensory and nociceptive input, and evidence of altered activity in this region in chronic low back pain^15^, targeting the sensorimotor cortex may provide a mechanistically informed approach to neuromodulation in CLBP. Although multi-band neurofeedback protocols have been explored in some chronic pain conditions^16–18^, they have not been evaluated in CLBP, nor have prior CLBP studies targeted the sensorimotor cortex. Accordingly, the in-house-developed neurofeedback system (PainWaive) used in this study was designed to (1) target multiple pain-related frequency bands and (2) record from C1 and C2 electrode sites located over the sensorimotor cortex. This digitally-delivered EEG neurofeedback involves recording surface EEG, extracting and processing the targeted frequency bands, and providing real-time feedback to individuals in a gamified interface based on their ongoing brain activity^19,20^. The goal of this intervention is to train individuals to modulate these specific frequency bands over the sensorimotor cortex to reduce pain severity.

This study employed a multiple-baseline single-case experimental design (SCED) to examine the potential effects of this digitally-delivered EEG neurofeedback intervention on pain severity and pain interference in individuals with CLBP. The study also evaluated the usability and acceptability of this novel system and explored whether the intervention influenced a variety of psychological outcomes, such as depression, anxiety, wellbeing, and sleep disturbance.

## METHODS

### Study design

This study used a SCED with multiple baselines across four participants to evaluate the effects of a home-based, self-directed EEG neurofeedback intervention for individuals with CLBP. Single-case designs provide an alternative to group-based designs when early-phase intervention testing is needed before scaling up to randomised controlled trials (RCTs)^21,22^. By relying on small, intensively monitored samples, SCEDs allow evaluation of intervention effects at the individual level, which is often obscured in group-based designs^22,23^.

The methodological rigour of a SCED is grounded in the repeated assessment of outcomes across at least two phases within the same participant^24^. In this study, each participant completed daily ratings of pain severity and pain interference across four phases: baseline, intervention, post-intervention, and follow-up. Because each participant’s baseline served as their own control condition, changes during and after the intervention were interpreted relative to baseline patterns. Continuous daily assessments helped detect intervention effects even in the presence of day-to-day variability in pain, which is often lost in group-level pre–post averages.

A multiple-baseline structure was used to strengthen internal validity and minimise the influence of time-related confounds. Baseline durations were staggered across participants, increasing sequentially to allow the intervention to begin at different time points. A minimum between-baseline difference of three days ensured that any improvement could be attributed to the intervention rather than temporal factors or personal events^25,26^. Demonstrating consistent changes following four distinct start-points provides strong evidence that observed intervention effects were linked to the introduction of neurofeedback rather than extraneous influences^22,25^.

This trial was conducted in accordance with best-practice SCED recommendations and guidelines^27,28^. Participants and the research team were not blinded to the phase allocations.

### Participants

Four participants were recruited based on the following eligibility criteria: (1) experiencing low back pain (with or without referred leg pain), defined as pain between the lower margin of the twelfth ribs to the lower gluteal folds^29^, (2) experiencing persistent pain for at least three months, (3) reporting an average pain severity of ≥4 on a 0–10 Numerical Rating Scale over the past seven days (4) aged 18–80 years, and (5) residing in Australia. Individuals were excluded if they were diagnosed with neurological or psychiatric disorders, or were expected to undergo changes in their current treatment regimen during the trial (a maximum of 12 weeks).

This study was approved by the University of New South Wales Human Research Ethics Committee (iRECS8134) and was registered in the Australian New Zealand Clinical Trials Registry (ACTRN12625000161426). Individuals with chronic low back pain were identified through an existing database at the NeuroRecovery Research Hub, UNSW, and were emailed to obtain consent for a telephone screening interview to assess their interest and eligibility for the study. All participants provided informed consent before the trial commenced.

### Procedure

#### Baseline phase (A)

All participants began the baseline phase on the same day. They were randomly assigned to one of four staggered baseline durations (7, 10, 14, or 17 days), ensuring a minimum of three assessment days difference between participants^27^. During this phase, participants provided daily ratings of the primary and secondary outcomes.

#### Intervention phase (B)

The intervention phase started immediately after each participant completed their assigned baseline period. Participants self-administered 20 daily sessions of EEG neurofeedback over a four-week period. Daily outcome ratings continued throughout this phase.

### EEG neurofeedback intervention

PainWaive, our in-house-developed EEG neurofeedback system, was used as the intervention in this study. The system includes three integrated components: (1) the PainWaive Gen-2 EEG headset, (2) the PainWaive web application, and (3) a secure cloud-based researcher dashboard.

Prior to the intervention phase, each participant received a home-based PainWaive kit^20^, containing the headset, a Surface Pro tablet preloaded with the web application, accessories, and a detailed user manual. Both the headset and the web application were designed and developed to support remote implementation of the digitally-delivered neurofeedback intervention. To ensure correct setup and familiarity with the procedures, participants completed their first two sessions via Zoom with a member of the research team for support. For the remaining sessions, the researcher dashboard enabled secure post-session monitoring of protocol adherence and EEG signal quality.

The PainWaive neurofeedback system (Figure 1) targets three frequency bands, aiming to suppress theta (θ, 4–7 Hz) and high-beta (high-β, 20–30 Hz) activity, while enhancing sensorimotor rhythms (SMR, 10–15 Hz). The headset records EEG from two electrodes positioned at C1 and C2 sites over the sensorimotor cortex, with two ear-clip electrodes serving as ground and reference. Signals were sampled at 200 Hz. Unlike most commercially available home-based EEG systems, the PainWaive headset incorporates impedance measurement to verify a good connection between the electrodes and the scalp.

**Figure 1.**
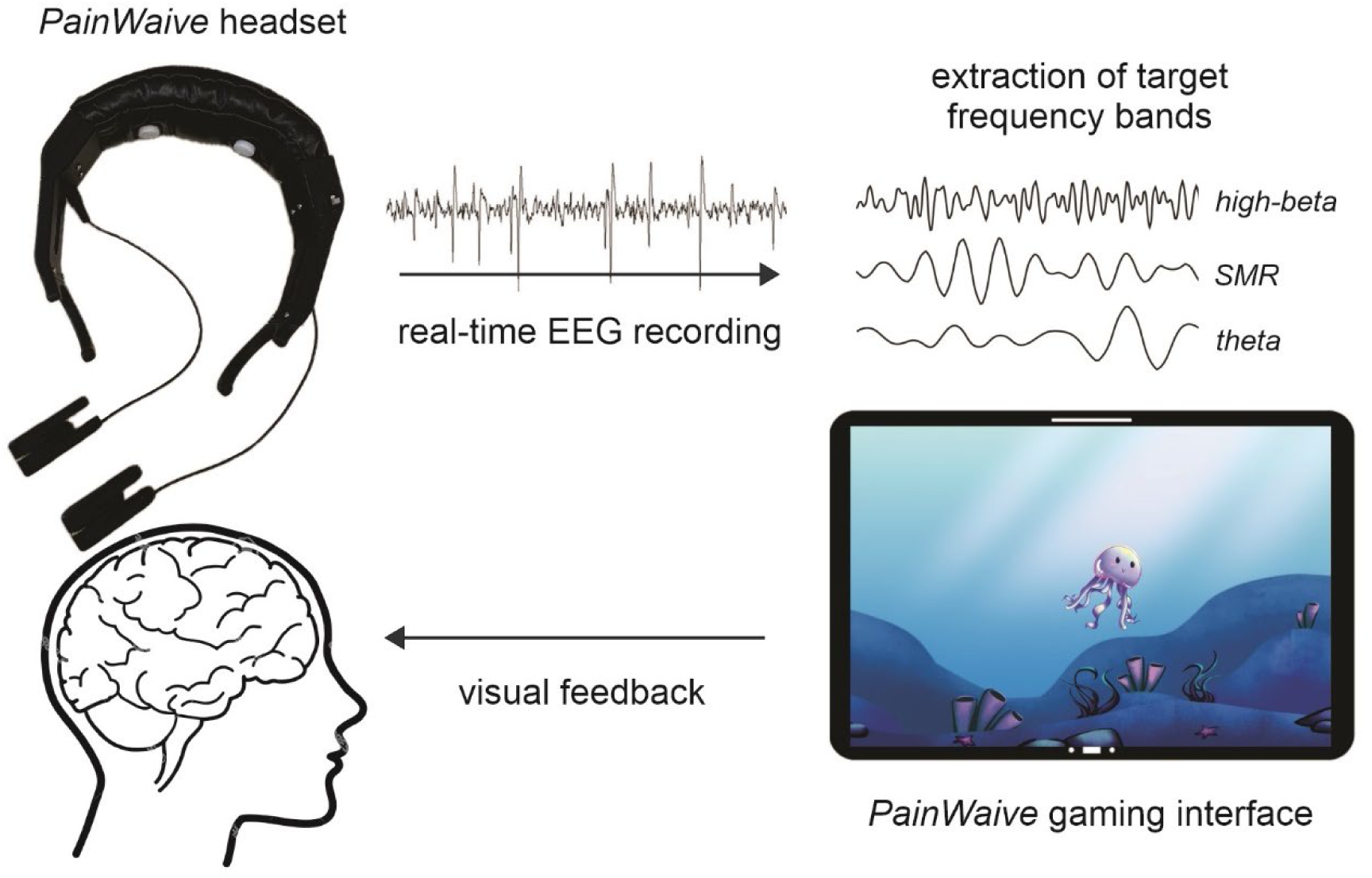
PainWaive neurofeedback schematic. Surface EEG is recorded via two sensors positioned over the sensorimotor cortex using the PainWaive headset; the targeted frequency bands are extracted and processed in real-time, and modulation of the band powers is presented to participants as positive visual feedback through the PainWaive gaming interface.

The web application provides step-by-step instructions for headset setup, impedance verification, resting-state EEG recording, and gameplay. Each session commenced with a photograph captured via the tablet to confirm correct headset placement, followed by impedance checks and a 2-minute eyes open recording, which served as the session’s resting-state threshold. Participants then completed five rounds of 2.5-minute neurofeedback game.

During gameplay, the application computed the relative power of the targeted frequency bands in real-time and compared these values to the resting-state threshold. Positive feedback was delivered when participants simultaneously decreased theta and high-beta rhythms while increasing SMR. Feedback was conveyed through changes in background colour and the movement of the game character within a gamified interface. No EEG data were displayed to participants. Four game scenarios (Jellyfish, Bird, Plane, Rocket) were rotated in blocks of five sessions to maintain engagement. At the end of each round, participants received a performance score reflecting the cumulative duration of successful neuromodulation. Background music was included to promote relaxation, with adjustable volume settings. Participants were encouraged to apply recommended mental strategies to reinforce learning associations between mental states and the game visual feedback.

All session data and headset placement photographs were automatically stored on the PainWaive secure dashboard, only accessible to the research team. The dashboard enabled monitoring of session completion, EEG signal quality, and headset placement. EEG signals were checked for any unexplained artifacts or noise, and participants were contacted with solutions, if necessary. Participants could also seek additional support via phone or videoconferencing throughout the trial.

#### Post-intervention and Follow-up phases (C and D)

Following the intervention, participants continued providing daily outcome ratings over two additional seven-day phases: one immediately after (post-intervention) and one five weeks later (follow-up). These phases were designed to evaluate the immediate and longer-term effects of the intervention. Participants were encouraged to recall the mental strategies used during the neurofeedback sessions and apply them in daily life to help consolidate their ability to regulate pain-related brain rhythms.

### Outcome measures

#### Primary outcome

The primary outcome was pain severity, assessed using the Pain Severity subscale of the Brief Pain Inventory (BPI)^30^. Participants rated their pain on a 0–10 Numerical Rating Scale (0 = “No pain”, 10 = “Worst imaginable pain”) across four domains: pain at the time of reporting, average pain over the past 24 hours, and the worst and least pain experienced over that period. Scores were averaged to generate a daily pain severity score, with higher scores indicating more severe pain. The BPI-Pain Severity subscale has demonstrated good internal reliability in chronic pain samples (*Cronbach’s α* = 0.85)^31^.

#### Secondary outcome

The secondary outcome was pain interference, assessed using the Pain Interference subscale of the BPI^30^. Participants rated how much pain interfered with seven aspects of daily life: general activity, normal work, walking ability, enjoyment of life, relationships with others, mood, and sleep, on a 0–10 Numerical Rating Scale (0 = “Does not interfere”, 10 = “Completely interferes”). These ratings were averaged to produce a daily pain interference score. The BPI-Pain Interference subscale has shown strong internal consistency (*Cronbach’s α* = 0.88)^31^.

#### Acceptability, usability, and perceived change

Participants took part in a semi-instructed Zoom interview the day after the intervention phase was completed. The interview invited them to reflect on their experiences with the trial procedures and the neurofeedback intervention. Interviews lasted approximately 40–50 minutes and were recorded with participants’ consent to minimise recall bias. Zoom transcripts were then manually checked for accuracy.

In addition, participants completed the System Usability Scale (SUS)^32^ and the Patient Global Impression of Change (PGIC)^33^ the day after the intervention phase. The SUS assessed perceived usability of the EEG neurofeedback system across 10 items rated on a 5-point Likert scale (1 = “Strongly disagree”, 5 = “Strongly agree”). Item scores were converted to a total score ranging 0– 100, with scores <50 indicating substantial usability problems and scores ≥85 indicating excellent usability^34^. The PGIC assessed participants’ perceived change in activity limitations, symptoms, emotions, and overall quality of life related to their pain condition since the start of treatment on a 7-point Likert scale (1 = no change/worse to 7 = a great deal better).

#### Generalisation measures

To determine whether any effects of the intervention extended beyond the daily-measured outcomes^21^, we assessed depressive symptoms, anxiety, wellbeing, and sleep disturbance at three timepoints. Beck Depression Inventory-II (BDI-II), the State-Trait Anxiety Inventory - State (STAI-S), COMPAS-Wellbeing scale, and PROMIS sleep disturbance short form were administered at baseline onset (T1), on the first day of post-intervention (T2), and on the first day of follow-up (T3).

The BDI-II assesses the severity of depressive symptoms, with scores than can range from 0 to 63^35^. Standard clinical cut-offs classify scores into minimal (0–13), mild (14–19), moderate (20–28), and severe (29–63) depression. This measure has demonstrated good test-retest reliability (*r* = 0.73–0.96)^36^. The STAI-S evaluates current (state) anxiety, with scores from 20 to 80^37^. As a state-sensitive measure, it does not have validated clinical cut-offs. Therefore commonly referenced severity bands (e.g., low: 20–37, moderate: 38–44, high: 45–80), derived from normative data^37,38^, were used to contextualise changes in anxiety. The wide range of test-retest reliability of this measure (*r* = 0.31–0.86) reflects the scale’s sensitivity to fluctuations in transient anxiety state^38^. The COMPAS-Wellbeing scale is used to measure total wellbeing with scores ranging from 26 to 130^39^. Resulting from normative samples, scores were categorised as languishing (≤89), moderate (90–110), or flourishing (≥111)^40^. This scale has demonstrated good test-retest reliability (*r* = 0.82)^39^. The PROMIS sleep disturbance short form yields scores ranging from 8 to 40. Raw scores are typically converted to T-scores (M = 50, SD = 10), which are interpreted as none to slight (<55), mild (55–59.9), moderate (60–69.9), and severe (≥70) sleep disturbance^41^. This measure has demonstrated high internal consistency (*Cronbach’s α* = 0.89–0.93)^42^.

### Data Analysis

#### Primary and secondary outcome measures

Participants’ daily ratings of pain severity and pain interference were compared between the baseline phase (A) and the intervention (B), post-intervention (C), and follow-up (D) phases. To evaluate whether there were any reliable intervention effect across phases, we used within-phase and between-phase analyses across two complementary approaches: visual inspection^24,43^ and *Tau-U* effect estimates^44^. Between-phase comparisons were between baseline and intervention (A vs B), baseline and post-intervention (A vs C), and baseline and follow-up (A vs D).

Visual inspection of graphs, with outcome data plotted across phases, involved examining the baseline stability, within-phase data patterns, and between-phase comparisons. The accuracy of visual analysis is strengthened when baseline data show relative stability. Baseline data was considered stable if ≥80% of datapoints fell within ±25% of the phase median^43^.

Within-phase visual analysis involved calculating the median, mean, and trend of the outcome data for each phase^43^. Trend reflects a systematic increase or decrease in the outcome data over time^24^, and was quantified using the split-middle approach^43^. In this study, a positive trend indicated an upward slope, contrary to the expected intervention effect, while a negative trend indicated a downward slope, consistent with the intended intervention effect.

Between-phase visual analysis evaluates whether changes in the outcomes correspond to the introduction of the intervention by examining four key features^24^. The first two are changes in the mean and median of pain severity and pain interference across phases. Although changes in these metrics alone may not be definitive, they contribute to the overall interpretation of potential intervention effects. The third feature is the relative change in level, which reflects any discontinuity between the second half of baseline phase and the first half of the intervention phase. A level change can capture a delayed effect following the introduction of the intervention phase and it may occur even without a corresponding change in mean. The fourth feature is the change in trend across phases, which can signal a systematic change in the data pattern attributable to the intervention.

*Tau-U* analysis is a widely used statistical method that complements visual inspection in SCEDs. It represents a set of non-parametric effect size indices that jointly capture both the trend within phases and the degree of change between phases^45^. Compared to other non-parametric effect size metrics^46^, *Tau-U* has shown superior discriminative ability and sensitivity, is less affected by autocorrelation, and allows controlling for an undesirable baseline trend^45,47^. *Tau-U* calculations were conducted using the online tool developed by Vannest and colleagues^48^. When a significant baseline trend was detected in the same direction as the expected intervention effect, baseline trend correction was applied. Individual-level *Tau-U* effect estimates were obtained and then aggregated to provide an overall intervention effect across all four participants. Intervention effects were interpreted using recommended thresholds: <0.20 indicating a small or negligible effect, 0.20–0.59 a medium effect, 0.60–0.79 a large effect, and ≥0.80 a very large effect^44^.

#### Acceptability, usability, and perceived change

Quotes related to the acceptability of the EEG neurofeedback intervention were extracted from the Zoom session transcripts and categorised using the components of the Theoretical Framework of Acceptability (TFA). In addition, all participants completed the SUS and PGIC, and the responses were reported for each participant.

#### Generalisation measures

The Reliable Change Index (RCI) was used to assess whether changes in the generalisation measures across the three timepoints (T1, T2, T3) represented reliable improvement or deterioration for each participant. RCI was evaluated for two intervals: T1 to T2 and T1 to T3. Calculations incorporated each participant’s score change, the normative standard deviation of the measure, and its test–retest reliability coefficient (or *Cronbach’s α* when test-retest coefficient was unavailable). An RCI exceeding ±1.96 indicates that the change is unlikely to be due to measurement errors and can therefore be considered reliable. In addition to the RCI, clinically relevant changes in the generalisation measures were evaluated based on established clinical cut-offs.

## RESULTS

### Participants’ characteristics

All four participants reported low back pain radiating down one or both legs during screening, with a severity of at least 4 (on a 0–10 scale) and duration exceeding three months. The PainDETECT questionnaire was administered to detect a neuropathic pain component. P1 obtained a score of 13, which falls within the ambiguous range, indicating that a neuropathic component may be present but is unclear. The remaining participants scored above 19, consistent with a likely neuropathic pain component according to PainDETECT classification criteria. Questionnaire scores and other participant characteristics are presented in Table 1.

**Table 1.** Participants’ characteristics at the time of screening.

|  | Age range | Sex (F/M) | Pain characteristics |  |  |  | Medications for pain relief *** |
| --- | --- | --- | --- | --- | --- | --- | --- |
|  |  |  | Duration of pain (y) | Severity of pain (0-10) * | PainDETECT score (0-38) ** | Location of pain |  |
| **P1** | 70-79 | F | 5 | 8 | 13 | Low back pain, radiating down the legs | <sup>1</sup> Lidocaine dermal patches<br><sup>2</sup> Diclofenac (Voltaren gel) |
| **P2** | 60-69 | M | 12 | 5 | 24 | Low back pain, radiating down the legs | <sup>3</sup> CBD oil with THC, 0.4 mL per day<br><sup>4</sup> Amitriptyline, 50 mg per day<br><sup>5</sup> Indomethacin suppositories |
| **P3** | 30-39 | M | 5.5 | 5 | 22 | Low back pain, radiating down the legs | None |
| **P4** | 70-79 | F | 20 | 4 | 21 | Low back pain, radiating down the legs | <sup>6</sup> Paracetamol, 665 mg per day<br><sup>2</sup> Diclofenac (Voltaren gel) |

P: Participant
\* Participants rated their pain severity on a 0-10 scale based on the average level of pain they experienced over the seven days preceding the screening day.
\*\* PainDETECT was used to screen for a neuropathic pain component. A score of $\geq 19$ suggests that a neuropathic pain component is likely, while scores of 13-18 are considered ambiguous, although neuropathic pain may be present.
\*\*\* Participants were asked to maintain their current medication regimen throughout the trial and to inform the Trial Research Team if any changes occurred.
<sup>1</sup> Lidocaine dermal patches: a topically applied local anaesthetic for reducing pain in the skin.
<sup>2</sup> Diclofenac: a topically applied non-steroidal anti-inflammatory drug (NSAID).
<sup>3</sup> CBD oil with THC: Cannabidiol (CBD) and Tetrahydrocannabinol (THC) are active compounds from the *Cannabis sativa* plant species (i.e., cannabinoids).
<sup>4</sup> Amitriptyline: a tricyclic antidepressant recommended for neuropathic pain.
<sup>5</sup> Indomethacin: a NSAID.

### Adherence

All four participants completed 20 intervention sessions within the 4-week period, and no adverse events were reported. Daily outcome ratings were maintained throughout, including non-intervention days. P4 completed a longer baseline phase (20 rather than 17 days) due to scheduling of the first supervised session, while still maintaining the required staggered design. During follow-up phases, P2 and P4 missed one and two assessments, respectively, but all phases met the minimum data points requirements. All participants completed the generalisation measures, PGIC and SUS questionnaires, and the semi-structured interview.

### Analysis of the primary outcome: pain severity

#### Visual inspection

Daily pain severity ratings for each participant were plotted for visual analysis (Figure 2), with median and split-middle trend lines overlaid for a clear interpretation across phases. The results of the within-phase and between-phase analyses are presented in Tables 2 and 3. Baseline data were stable across all participants, with no notable trend except for P1, who showed an upward (contra-therapeutic) trend during baseline.

**Figure 2.**
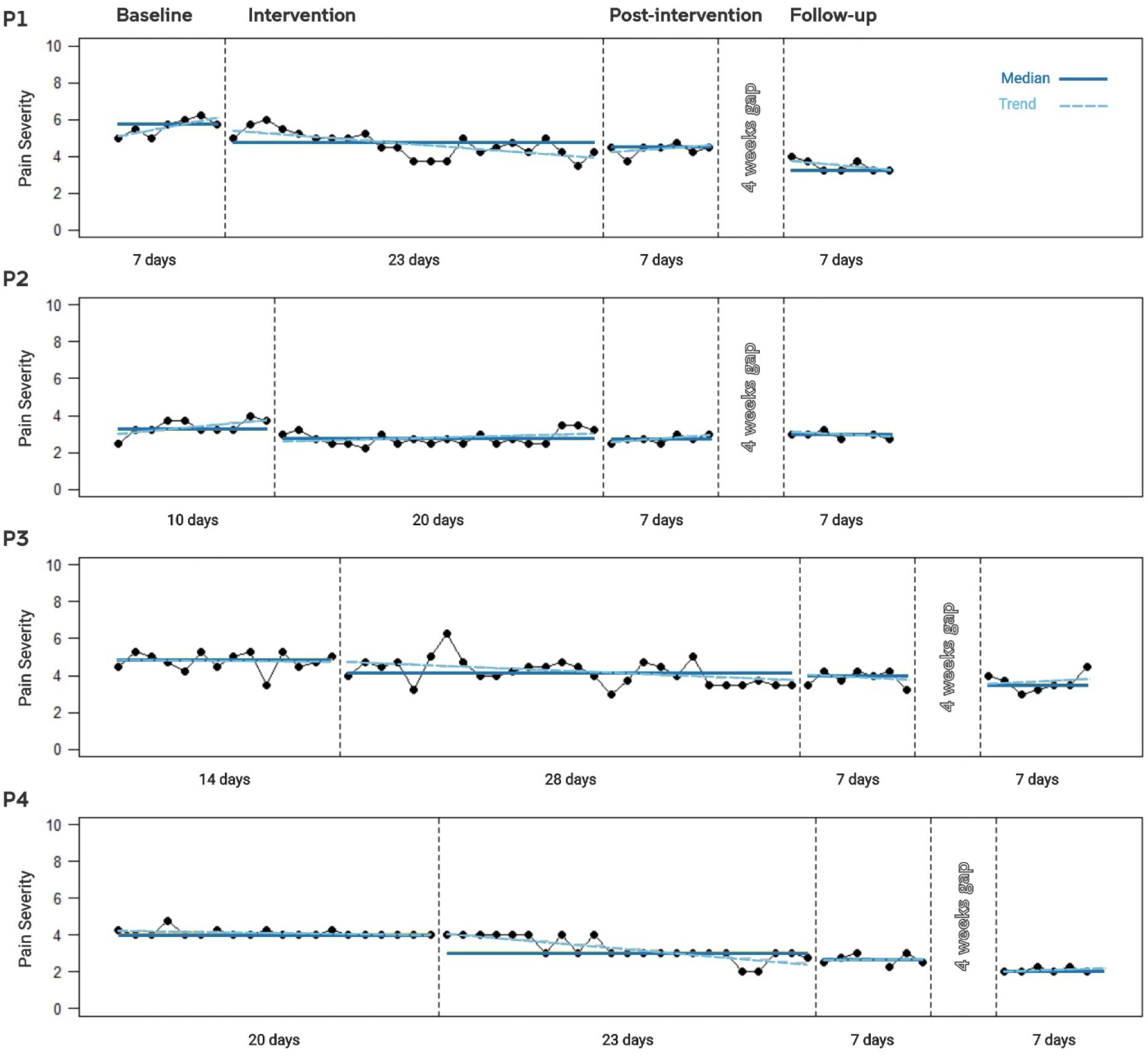
Individuals’ pain severity across all phases of the trial. Pain severity is plotted for each participant (P1-P4) throughout the baseline, intervention, post-intervention, and follow-up phases. Solid lines indicate phase medians, and dashed lines represent the within-phase trend. Participants completed 20 sessions over four weeks, and daily ratings were recorded on both intervention and non-intervention days; therefore, the number of datapoints during the intervention phase varies across participants.

**Table 2.** Within-phase analysis of pain severity data for all four participants. Median, mean, and trend are reported for each phase, and stability is reported for the baseline phase.

| **Within-phase analysis – Pain Severity** |  |  |  |  |  |  |  |  |  |  |  |  |  |
| --- | --- | --- | --- | --- | --- | --- | --- | --- | --- | --- | --- | --- | --- |
|  | **Baseline phase (A)** |  |  |  | **Intervention phase (B)** |  |  | **Post-intervention phase (C)** |  |  | **Follow-up phase (D)** |  |  |
|  | Median | Mean | Trend | Stability * | Median | Mean | Trend | Median | Mean | Trend | Median | Mean | Trend |
| **P1** | 5.75 | 5.61 | 0.25 | 100% | 4.75 | 4.68 | -0.06 | 4.50 | 4.39 | 0.00 | 3.25 | 3.50 | -0.14 |
| **P2** | 3.25 | 3.40 | 0.00 | 100% | 2.75 | 2.79 | 0.01 | 2.75 | 2.75 | 0.04 | 3.00 | 2.96 | -0.08 |
| **P3** | 4.88 | 4.77 | 0.04 | 93% | 4.13 | 4.21 | -0.05 | 4.00 | 3.89 | 0.11 | 3.50 | 3.64 | -0.07 |
| **P4** | 4.09 | 4.00 | 0.00 | 100% | 3.25 | 3.00 | -0.09 | 2.67 | 2.63 | -0.08 | 2.08 | 2.00 | 0.00 |

\* Percentage of scores within the stability envelope ( $\pm 25\%$ of the median).

**Table 3.** Between-phase analysis of pain severity data for all four participants. Changes in mean, median, and trend are reported when comparing baseline (A) phase with intervention (B), post-intervention (C), and follow-up (D) phases. Relative change in level is reported for the comparison between baseline (A) and intervention (B).

| **Between-phase analysis – Pain Severity** |  |  |  |  |  |  |  |  |  |  |
| --- | --- | --- | --- | --- | --- | --- | --- | --- | --- | --- |
|  | **Change in median** |  |  | **Change in mean** |  |  | **Change in trend** |  |  | **Relative change in level \*** |
|  | **A vs B** | **A vs C** | **A vs D** | **A vs B** | **A vs C** | **A vs D** | **A vs B** | **A vs C** | **A vs D** | **A vs B** |
| **P1** | -1.00 | -1.25 | -2.50 | -0.92 | -1.21 | -2.11 | -0.31 | -0.25 | -0.39 | -1.00 |
| **P2** | -0.50 | -0.50 | -0.25 | -0.61 | -0.65 | -0.44 | 0.01 | 0.04 | -0.08 | -0.63 |
| **P3** | -0.75 | -0.88 | -1.83 | -0.55 | -0.88 | -1.13 | -0.09 | 0.07 | -0.11 | -0.50 |
| **P4** | -1.00 | -1.38 | -2.00 | -0.84 | -1.42 | -2.00 | -0.09 | -0.08 | 0.00 | 0.00 |

\* Relative change in level is defined as the change in median from the second half of one phase to the first half of the adjacent phase; therefore, it is reported only for the comparison between baseline (A) and intervention (B).

Inspection of changes in median and mean values indicated that P1, P3, and P4 had consistent, phase-by-phase reductions in pain severity, while P2 showed less consistent and minimal pain reductions across phases. For the three of four participants, the most pronounced improvements emerged when comparing baseline to the follow-up phase, with reductions ranging from 1.1 to 2.5 points. Relative change in level, defined as the shift in median from the latter half of baseline to the initial half of the intervention phase, suggested that improvements during intervention occurred with latency in P3 and P4.

#### Statistical analysis

The within-phase trend estimates from the *Tau* analysis confirmed the presence of an upward baseline trend for P1 (Table 4). Since this trend was in the contra-therapeutic direction, baseline correction was not required. Individual-level *Tau-U* effect estimates are reported in Table 5 for all comparisons. An aggregated *Tau-U* effect estimate was also calculated across all four participants (Table 5). This analysis indicated a large effect when comparing baseline to intervention (*Tau* = - 0.67, 95% CI [-0.88, −0.46]), and very large effects for both baseline to post-intervention (*Tau* = - 0.92, 95% CI [-1, −0.63]) and baseline to follow-up comparisons (*Tau* = −0.90, 95% CI [-1, −0.61]).

**Table 4.** Within-phase trends in pain severity for each participant, calculated using *Tau* analysis across baseline (A), intervention (B), post-intervention (C), and follow-up (D) phases. 90% confidence intervals (CIs) are reported; CIs not including zero indicate a significant trend (shown in bold).

| ***Tau trend calculation – Pain Severity*** |  |  |  |  |  |  |  |  |
| --- | --- | --- | --- | --- | --- | --- | --- | --- |
|  | ***Trend A*** |  | ***Trend B*** |  | ***Trend C*** |  | ***Trend D*** |  |
|  | ***Tau*** | ***90% CI*** | ***Tau*** | ***90% CI*** | ***Tau*** | ***90% CI*** | ***Tau*** | ***90% CI*** |
| **P1** | **0.65** | **[0.10, 1]** | **-0.55** | **[-0.76, -0.27]** | 0.17 | [-0.38, 0.66] | -0.57 | [-1, 0.05] |
| **P2** | 0.47 | [-0.01, 0.81] | 0.12 | [-0.16, 0.38] | 0.54 | [-0.05, 1] | -0.38 | [-0.92, 0.25] |
| **P3** | 0 | [-0.33, 0.33] | **-0.34** | **[-0.54, -0.10]** | 0 | [-0.52, 0.52] | 0.10 | [-0.43, 0.62] |
| **P4** | -0.26 | [-0.45, 0.08] | **-0.68** | **[-0.80, -0.31]** | 0.07 | [-0.52, 0.65] | 0.17 | [-0.45, 0.72] |

**Table 5.** *Tau-U* effect size for pain severity in each participant, comparing baseline to intervention (*A vs B*), baseline to post-intervention (*A vs C*), and baseline to follow-up (*A vs D*). Overall effect size across all four participants is also reported. Confidence intervals (CIs) are reported; CIs not including zero indicate significant effect sizes (shown in bold).

| ***Tau-U effect size – Pain Severity*** |  |  |  |  |  |  |
| --- | --- | --- | --- | --- | --- | --- |
|  | ***A vs B*** |  | ***A vs C*** |  | ***A vs D*** |  |
|  | ***Tau*** | ***90% CI*** | ***Tau*** | ***90% CI*** | ***Tau*** | ***90% CI*** |
| **P1** | **-0.76** | **[-1, -0.31]** | **-1** | **[-1, -0.47]** | **-1** | **[-1, -0.47]** |
| **P2** | **-0.73** | **[-1, -0.33]** | **-0.84** | **[-1, -0.35]** | **-0.75** | **[-1, -0.21]** |
| **P3** | **-0.57** | **[-0.86, -0.23]** | **-0.87** | **[-1, -0.41]** | **-0.89** | **[-1, -0.42]** |
| **P4** | **-0.85** | **[-1, -0.44]** | **-1** | **[-1, -0.55]** | **-1** | **[-1, -0.55]** |
| **Weighted average** | ***Tau*** | ***95% CI*** | ***Tau*** | ***95% CI*** | ***Tau*** | ***95% CI*** |
|  | **-0.67** | **[-0.88, -0.46]** | **-0.92** | **[-1, -0.63]** | **-0.90** | **[-1, -0.61]** |

### Analysis of the secondary outcome: pain interference

#### Visual inspection

Daily pain interference data were also plotted for each participant (Figure 3), with median and trend lines included to assist interpretation across phases. The within-phase and between-phase visual analyses are summarised in Tables 6 and 7. Pain interference data were stable during baseline for all except P3, who showed fluctuations across the 14-day baseline. However, these fluctuations were not reflected in the split-middle trend analyses. No notable baseline trend was observed across participants.

**Figure 3.**
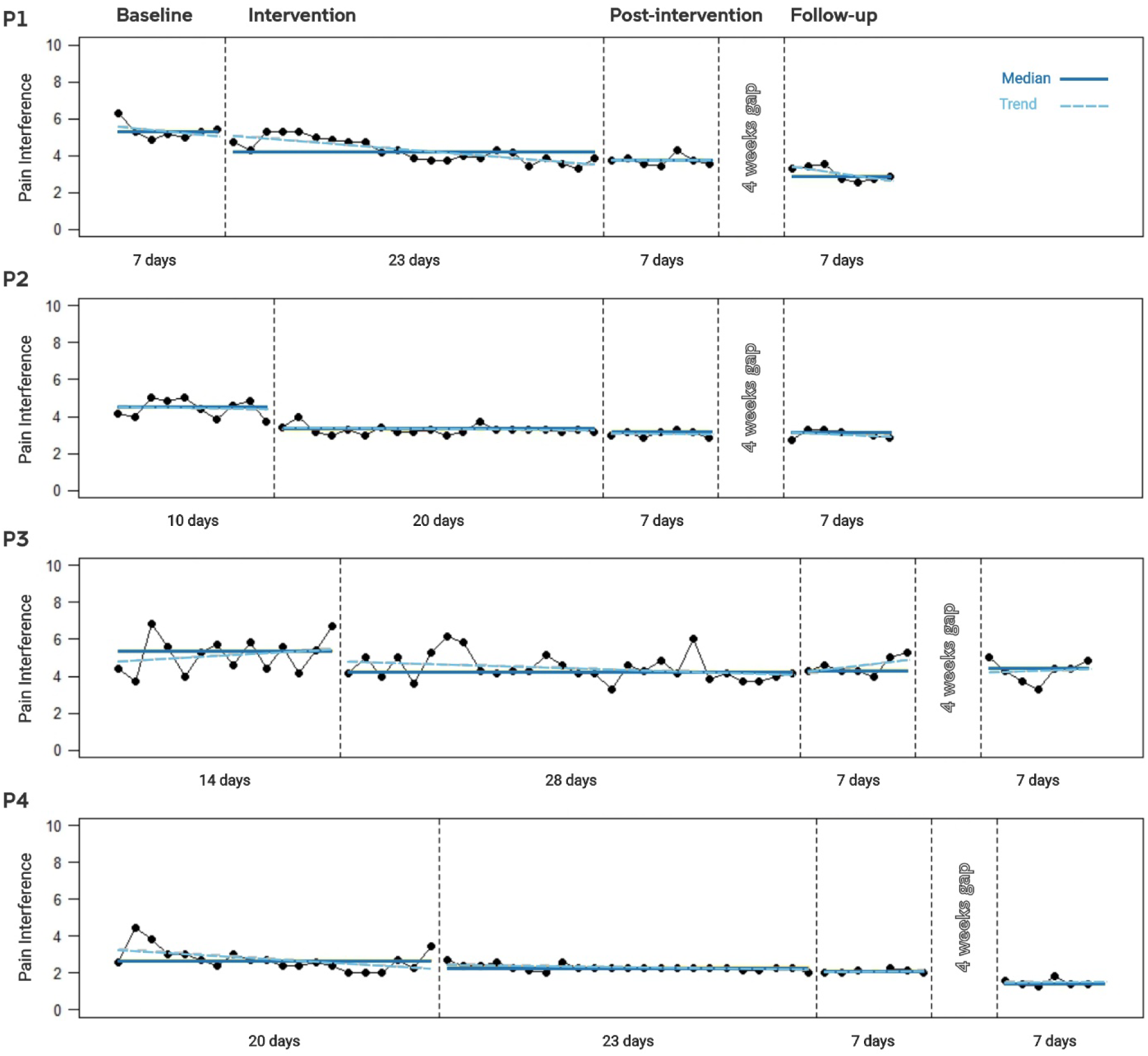
Individuals’ pain interference across all phases of the trial. Pain interference is plotted for each participant (P1-P4) throughout the baseline, intervention, post-intervention, and follow-up phases. Solid lines indicate phase medians, and dashed lines represent the within-phase trend. Participants completed 20 sessions over four weeks, and daily ratings were recorded on both intervention and non-intervention days; therefore, the number of datapoints during the intervention phase varies across participants.

**Table 6.** Within-phase analysis of pain interference data for all four participants. Median, mean, and trend are reported for each phase, and stability is reported for the baseline phase.

| **Within-phase analysis – Pain Interference** |  |  |  |  |  |  |  |  |  |  |  |  |  |
| --- | --- | --- | --- | --- | --- | --- | --- | --- | --- | --- | --- | --- | --- |
|  | **Baseline phase (A)** |  |  |  | **Intervention phase (B)** |  |  | **Follow-up I phase (C)** |  |  | **Follow-up II phase (D)** |  |  |
|  | Median | Mean | Trend | Stability * | Median | Mean | Trend | Median | Mean | Trend | Median | Mean | Trend |
| **P1** | 5.29 | 5.33 | -0.02 | 100% | 4.14 | 4.27 | -0.07 | 3.71 | 3.73 | -0.02 | 2.86 | 3.02 | -0.21 |
| **P2** | 4.50 | 4.44 | -0.09 | 100% | 3.29 | 3.27 | 0.01 | 3.14 | 3.06 | 0.04 | 3.07 | 3.05 | -0.10 |
| **P3** | 5.36 | 5.16 | 0.02 | 71% | 4.21 | 4.45 | -0.02 | 4.29 | 4.53 | 0.10 | 4.43 | 4.29 | 0.04 |
| **P4** | 2.64 | 2.74 | -0.04 | 85% | 2.29 | 2.30 | 0.00 | 2.07 | 2.10 | 0.05 | 1.43 | 1.50 | 0.00 |

\* Percentage of scores within the stability envelope ( $\pm 25\%$ of the median).

**Table 7.** Between-phase analysis of pain interference data for all four participants. Changes in mean, median, and trend are reported when comparing baseline (A) phase with intervention (B), post-intervention (C), and follow-up (D) phases. Relative change in level is reported for the comparison between baseline (A) and intervention (B).

| **Between-phase analysis – Pain Interference** |  |  |  |  |  |  |  |  |  |  |
| --- | --- | --- | --- | --- | --- | --- | --- | --- | --- | --- |
|  | **Change in median** |  |  | **Change in mean** |  |  | **Change in trend** |  |  | **Relative change in level \*** |
|  | **A vs B** | **A vs C** | **A vs D** | **A vs B** | **A vs C** | **A vs D** | **A vs B** | **A vs C** | **A vs D** | **A vs B** |
| **P1** | -1.14 | -1.57 | -2.43 | -1.06 | -1.59 | -2.31 | -0.05 | 0.00 | -0.19 | -0.57 |
| **P2** | -1.21 | -1.36 | -1.43 | -1.17 | -1.38 | -1.40 | 0.09 | 0.13 | -0.01 | -1.21 |
| **P3** | -1.14 | -1.07 | -0.93 | -0.71 | -0.63 | -0.88 | -0.04 | 0.08 | 0.02 | -1.00 |
| **P4** | -0.36 | -0.57 | -1.21 | -0.44 | -0.64 | -1.24 | 0.04 | 0.09 | 0.04 | -0.14 |

\* Relative change in level is defined as the change in median from the second half of one phase to the first half of the adjacent phase; therefore, it is reported only for the comparison between baseline (A) and intervention (B).

Based on median and mean changes, P1, P2, and P4 demonstrated consistent phase-by-phase improvement in pain interference, while P3 showed less consistent reductions. For the three of four participants, the greatest improvements were observed between baseline and follow-up, with reductions ranging from 1.2 to 2.4 points. Relative change in level suggested a delayed onset of improvement during intervention for P1 and P4.

#### Statistical analysis

The within-phase trend estimates from the *Tau* analysis for pain interference identified a downward trend in baseline for P4 (Table 8). Because this trend was in the therapeutic direction, baseline trend correction was applied before calculating the overall *Tau-U* across all participants. Table 9 details the *Tau-U* values (with and without the baseline trend control) for each participant and all phase comparisons. The aggregated *Tau-U* effect estimates (Table 9) indicated large effects for all three comparisons: baseline to intervention (*Tau* = −0.63, 95% CI [-0.84, −0.41]), baseline to post-intervention (*Tau* = −0.62, 95% CI [-0.90, −0.33]), and baseline to follow-up (*Tau* = −0.70, 95% CI [-0.99, −0.41]).

**Table 8.** Within-phase trends in pain interference for each participant, calculated using *Tau* analysis across the baseline (A), intervention (B), post-intervention (C), and follow-up (D) phases. 90% confidence intervals (CIs) are reported, with CIs not including zero indicating significant trend (shown in bold).

| ***Tau trend calculation – Pain Interference*** |  |  |  |  |  |  |  |  |
| --- | --- | --- | --- | --- | --- | --- | --- | --- |
|  | ***Trend A*** |  | ***Trend B*** |  | ***Trend C*** |  | ***Trend D*** |  |
|  | ***Tau*** | ***90% CI*** | ***Tau*** | ***90% CI*** | ***Tau*** | ***90% CI*** | ***Tau*** | ***90% CI*** |
| **P1** | 0 | [-0.52, 0.52] | **-0.65** | **[-0.88, -0.39]** | -0.15 | [-0.66, 0.38] | -0.29 | [-0.81, 0.24] |
| **P2** | -0.16 | [-0.56, 0.25] | -0.03 | [-0.30, 0.23] | 0.05 | [-0.47, 0.57] | -0.28 | [-0.85, 0.32] |
| **P3** | 0.17 | [-0.16, 0.49] | **-0.24** | **[-0.46, -0.02]** | 0.31 | [-0.24, 0.81] | 0.10 | [-0.43, 0.62] |
| **P4** | **-0.46** | **[-0.70, -0.17]** | **-0.37** | **[-0.62, -0.13]** | 0.23 | [-0.38, 0.78] | -0.15 | [-0.72, 0.45] |

**Table 9.** *Tau-U* effect size for pain interference in each participant before and after controlling for the baseline trend, comparing baseline to intervention (*A vs B*), baseline to post-intervention (*A vs C*), and baseline to follow-up (*A vs D*). The overall effect size across all four is also reported. Confidence intervals (CIs) are reported, with CIs not including zero indicating significant effect sizes (shown in bold).

| ***Tau-U effect size – Pain Interference*** |  |  |  |  |  |  |
| --- | --- | --- | --- | --- | --- | --- |
|  | ***A vs B*** |  | ***A vs C*** |  | ***A vs D*** |  |
|  | ***Tau*** | ***90% CI*** | ***Tau*** | ***90% CI*** | ***Tau*** | ***90% CI*** |
| **P1** | **-0.85** | **[-1, -0.41]** | **-1** | **[-1, -0.47]** | **-1** | **[-1, -0.47]** |
| **P2** | **-0.97** | **[-1, -0.60]** | **-1** | **[-1, -0.52]** | **-1** | **[-1, -0.49]** |
| **P3** | **-0.44** | **[-0.75, -0.12]** | -0.42 | [-0.87, 0.03] | **-0.53** | **[-0.97, -0.07]** |
| **P4** | **-0.59** | **[-0.86, -0.27]** | **-0.80** | **[-1, -0.32]** | **-1** | **[-1, -0.55]** |

|  | ***A vs B - trend A*** |  | ***A vs C - trend A*** |  | ***A vs D - trend A*** |  |
| --- | --- | --- | --- | --- | --- | --- |
|  | ***Tau*** | ***90% CI*** | ***Tau*** | ***90% CI*** | ***Tau*** | ***90% CI*** |
| **P1** | **-0.85** | **[-1, -0.41]** | **-1** | **[-1, -0.41]** | **-1** | **[-1, -0.47]** |
| **P2** | **-0.94** | **[-1, -0.56]** | **-0.90** | **[-1, -0.56]** | **-0.88** | **[-1, -0.38]** |
| **P3** | **-0.48** | **[-0.78, -0.15]** | **-0.58** | **[-1, -0.41]** | **-0.69** | **[-1, -0.22]** |
| **P4** | **-0.40** | **[-0.68, -0.09]** | **-0.08** | **[-1, -0.56]** | **-0.31** | **[-0.76, 0.14]** |
| Weighted average | ***Tau*** | ***95% CI*** | ***Tau*** | ***95% CI*** | ***Tau*** | ***95% CI*** |
|  | **-0.63** | **[-0.84, -0.41]** | **-0.62** | **[-0.90, -0.33]** | **-0.70** | **[-0.99, -0.41]** |

### Acceptability, usability, and perceived change

The semi-structured interviews provided insights about the acceptability of the neurofeedback intervention. Acceptability was facilitated by participants’ confidence in self-administering the system, their understanding of the neurofeedback mechanism for pain management, and the perception that the intervention had achieved its intended purpose. The home-based delivery was consistently identified as a key facilitator, allowing flexibility and accessibility that would not have been feasible with daily clinic visits. After initial familiarisation, participants described the system as easy to set up and operate, supported by clear instructions and intuitive design. They expressed confidence in independent use, reported applying learned mental strategies outside training sessions, and indicated willingness to continue the intervention or recommend it to others with chronic pain. Reported barriers were relatively minor and included reduced motivation on workdays and on days with higher pain intensity, as well as challenges positioning the headset when wearing glasses. However, these issues did not appear to compromise engagement.

These qualitative findings were reflected in the quantitative usability ratings. The usability of the neurofeedback system was rated as good-to-excellent across all four participants on the SUS, with total scores of 85 (P1), 75 (P2), 87.5 (P3), and 95 (P4). Participants’ global impressions of change (PGIC) were also generally favourable: P1 and P2 reported being ‘*moderately better, with a slight but noticeable change*’, P3 reported being ‘*a little better, with no noticeable change*’, and P4 reported being ‘*better, with a definite improvement that has made a real and worthwhile difference*’.

### Generalisation measures

The generalisation measures assessed at three timepoints (T1, T2, and T3), along with the RCI values and clinically relevant changes, are reported in Table 10 for all four participants. The normative standard deviations and the reliability coefficients to calculate RCI were derived from the relevant literature^36,38–40,42,49–51^.

**Table 10.** The reliable change index and clinically relevant change for the generalisation measures across all four participants.

|  | Timepoints |  |  | SD of normative sample<sup>x</sup> | Reliability Coefficient<sup>o</sup> | Reliable Change Index |  |  |  | Clinically Relevant Change |  |
| --- | --- | --- | --- | --- | --- | --- | --- | --- | --- | --- | --- |
|  | T1 | T2 | T3 |  |  | T2-T1 |  | T3-T1 |  | T2-T1 | T3-T1 |
| BDI-II |  |  |  |  |  |  |  |  |  |  |  |
| P1 | 16 | 2 | 5 | 11.6 | 0.89 | -2.57 | Improved | -2.02 | Improved | Improved | Improved |
| P2 | 19 | 3 | 2 |  |  | -2.94 | Improved | -3.12 | Improved | Improved | Improved |
| P3 | 18 | 23 | 33 |  |  | 0.92 | NR | 2.76 | Worsened | Worsened | Worsened |
| P4 | 9 | 1 | 2 |  |  | -1.47 | NR | -1.29 | NR | NC | NC |
| STAI-S |  |  |  |  |  |  |  |  |  |  |  |
| P1 | 50 | 36 | 45 | 13.0 | 0.59 | -1.19 | NR | -0.42 | NR | Improved | NC |
| P2 | 45 | 34 | 29 |  |  | -0.93 | NR | -1.36 | NR | Improved | Improved |
| P3 | 49 | 62 | 69 |  |  | 1.10 | NR | 1.70 | NR | NC | NC |
| P4 | 43 | 25 | 31 |  |  | -1.53 | NR | -1.02 | NR | Improved | Improved |
| COMPAS-Wellbeing |  |  |  |  |  |  |  |  |  |  |  |
| P1 | 81 | 100 | 93 | 10.5 | 0.84 | 3.20 | Improved | 2.02 | Improved | Improved | Improved |
| P2 | 69 | 97 | 89 |  |  | 4.71 | Improved | 3.37 | Improved | Improved | NC |
| P3 | 56 | 63 | 61 |  |  | 1.18 | NR | 0.84 | NR | NC | NC |
| P4 | 90 | 106 | 110 |  |  | 2.69 | Improved | 3.37 | Improved | NC | NC |
| PROMIS sleep disturbance (t-score) |  |  |  |  |  |  |  |  |  |  |  |
| P1 | 56.3 | 47.9 | 54.3 | 10 | 0.91 | -1.98 | Improved | -0.47 | NR | Improved | Improved |
| P2 | 57.3 | 52.2 | 53.3 |  |  | -1.20 | NR | -0.94 | NR | Improved | Improved |
| P3 | 55.3 | 52.2 | 61.5 |  |  | -0.73 | NR | 1.46 | NR | Improved | Worsened |
| P4 | 49 | 44.2 | 44.2 |  |  | -1.13 | NR | -1.13 | NR | NC | NC |

**T1:** baseline onset, **T2:** post-intervention first day, **T3:** follow-up first day, **SD:** standard deviation, **BDI-II:** Beck Depression Inventory-II, **STAI-S:** State-Trait Anxiety Index-State, **NR:** not reliable, **NC:** no change.
<sup>x</sup> Standard deviations of normative samples were derived from the literature for each of the generalisation measures <sup>40,49-51</sup>.
<sup>o</sup> The reliability coefficients were drawn from psychometric studies. For the BDI-II, the reliability coefficient ranged from 0.84 to 0.95 <sup>36</sup>, the midpoint (0.89) was used here. For the STAI-S, a reliability ranged from 0.31 to 0.86 <sup>38</sup>, the midpoint (0.59) was used here. For the COMPAS-Wellbeing scale, a reliability of 0.84 <sup>39</sup> was used. Finally, reliability coefficients for the PROMIS sleep disturbance ranged from 0.89 to 0.93 <sup>42</sup>, and the midpoint (0.91) was used for the RCI analysis.

For depressive symptoms, reliable and clinically relevant changes were largely in agreement. P1 and P2 both had mild depressive symptoms at T1 and demonstrated reliable and clinically meaningful improvements by T2 that were maintained at T3, shifting toward the minimal depression range. P3 also reported mild depressive symptoms at T1 but showed reliable and clinically meaningful worsening by T2, with further deterioration by T3. P4 had minimal depressive symptoms at T1 and showed no reliable or clinically relevant change across timepoints.

For state anxiety, reliable and clinically relevant changes were not consistently aligned. No participant demonstrated reliable change across timepoints. Nevertheless, clinically relevant improvements from T1 to T2 were observed for P1, P2, and P4. By T3, P2 and P4 maintained these clinically relevant improvements, whereas P1 returned to baseline levels. P3 showed no clinically relevant change at any timepoint.

For wellbeing, reliable and clinically relevant changes were partially aligned. P1, P2, and P4 demonstrated reliable improvements across timepoints; however, clinically relevant improvement was observed only for P1 and P2, while P4 showed reliable improvement without corresponding clinical categorisation. P3 showed no reliable or clinically relevant change.

For sleep disturbance, reliable change and clinically relevant change were not aligned, with almost all changes not exceeding RCI threshold. However, P1 and P2 demonstrated clinically relevant improvements from T1 to T2 that were maintained at T3. P3 improved from T1 to T2, but showed clinically relevant worsening by T3, while P4 showed no reliable or clinically relevant change.

## DISCUSSION

In this trial, we explored the effects of the PainWaive EEG neurofeedback intervention in four individuals with chronic low back pain. The findings suggest that this digitally delivered intervention improves both pain severity and pain interference, with reductions observed across all four participants when comparing baseline to intervention, post-intervention, and follow-up phases. This remote intervention was deemed usable and acceptable by all participants. However, changes in psychological outcomes of anxiety, depression, wellbeing and sleep problems were mixed and varied across participants.

For pain severity, the aggregated *Tau-U* estimates indicated a large effect from baseline to intervention (*Tau-U* = −0.67) and very large from baseline to post-intervention and follow-up phases (*Tau-U* = −0.92 and −0.90). Individual effect estimates similarly suggested substantial reductions across participants. Visual inspection supported these findings: three participants (P1, P3, P4) demonstrated clear, progressive decreases in pain severity across phases, with the most pronounced improvements occurring from baseline to follow-up (reductions of 1.1–2.5 points), and P2 showed minimal and inconsistent change. Pain interference demonstrated a similarly favourable pattern. Aggregated *Tau-U* estimates indicated large effects for all comparisons (*Tau-U* = −0.63, −0.62, and −0.70), and individual effect estimates showed reductions for all participants except the baseline to post-intervention comparison for P3. Visual inspection revealed consistent phase-by-phase improvements for three participants (P1, P2, P4), with the greatest reductions again observed between baseline and follow-up (1.2–2.4 points). P3 displayed more variable trajectories over time, consistent with less robust statistical effects.

The magnitude of pain reduction observed in the present study appears at least comparable to, but in some instances larger than those reported in previous CLBP neurofeedback trials, although the methodological differences should be considered. Mayaud et al.^6^, in an open-label pilot of 20 alpha neurofeedback sessions, described broad clinical improvement; however, no clear statistically significant reductions in pain intensity were reported. Similarly, Shimizu et al.^8^ reported small and non-significant pre–post effects on pain [*d* = 0.22 in early CLBP (n=13) and *d* = 0.04 in late CLBP (n=7)], suggesting minimal analgesic effect of their protocol. In contrast, Adhia et al.^7^, in a four-arm RCT (n=15 per group), reported that their most responsive neurofeedback arm (targeting Infraslow frequency in pgACC) demonstrated mean reductions of −1.9 (95% CI: −2.7 to −1.0) in pain severity and −2.3 (95% CI: −3.5 to −1.2) in pain interference at one-month follow-up. These magnitudes are closely aligned with the 1.1–2.5-point reductions in pain severity and 1.2–2.4-point reductions in pain interference observed in three of four participants at follow-up in the present study. Notably, the values reported by Adhia et al. reflect within-group change rather than a comparison with placebo; therefore, both sets of findings represent improvements over time within an active neurofeedback condition. Although the RCT design permits between-group evaluation of intervention-specific effects, the multiple-baseline SCED used here provides intensive individual-level measurement and clearer temporal linkage between intervention introduction and outcome change. Also, the *Tau-U* estimates were large in the present study, this metric reflects non-overlap between phases rather than a standardised mean difference. It should therefore not be directly compared with effect sizes reported in other designs. Differences in neurofeedback protocols (e.g., targeting single-band versus multi-band frequency), session structures, and analytic approaches may also contribute to variation in observed effect sizes.

Beyond pain outcomes, psychological responses varied across participants. Consistent with prior reviews^9,52^, these findings suggest that EEG neurofeedback may produce more consistent effects on pain-related than on psychological outcomes. The variability observed across participants likely reflects individual differences rather than a uniform treatment effect and may relate to individual characteristics, and the extent of pain improvement. Given that the PainWaive neurofeedback protocol was designed to target pain-related brain rhythms, any psychological benefits are likely indirect and participant-specific.

The usability and acceptability findings were consistent with our previous SCED using the same neurofeedback protocol in corneal neuropathic pain^18^. Demonstrating similar implementation outcomes across two different pain conditions and two generations of the PainWaive headset increases confidence in the robustness of this digitally-delivered intervention, rather than suggesting condition-specific feasibility. The home setting may be particularly beneficial for individuals with chronic low back pain, who may face mobility limitations or practical barriers to attend daily clinic sessions. Furthermore, participants’ global impressions of change (PGIC) broadly aligned with observed pain reductions and reflected patterns seen across psychological outcomes, highlighting the value of integrating subjective experience with quantitative metrics. Taken together, the replication of favourable implementation outcomes and the convergence of subjective and objective findings illustrate how multiple-baseline experimental designs can provide rigorous early-phase evidence for both clinical and real-world feasibility prior to large-scale trials.

### Limitations and Strengths

Several methodological limitations should be considered when interpreting the findings of this study. Using the Risk of Bias in N-of-1 Trials (RoBiNT) scale^53^, this SCED achieved a total score of 21/30, with strong external validity (14/16), but more limited internal validity (7/14). The reduced internal validity was primarily due to the absence of blinding for participants, researchers, and assessors, as well as reliance on self-report outcomes, which prevented assessment of inter-rater agreement. Blinding was inherently not feasible in this design, constraining the level of internal control that could be achieved. The absence of a control condition further limits causal interpretation, as placebo effects and expectancy-related improvements cannot be ruled out, particularly given evidence that sham neurofeedback can produce subjective pain relief^54^. Self-administration of the intervention introduces additional sources of variability, including headset placement and environmental distractions. However, EEG signal quality and headset placement were monitored through the PainWaive dashboard and feedback was provided, when necessary.

Despite the limitations, this study has several strengths. The strong external validity reflects the real-world applicability of this digitally delivered intervention, which may enhance accessibility and adherence, particularly for individuals with mobility constraints or limited access to specialised care. The monitoring procedures and personalised feedback supported PainWaive intervention integrity and participants’ adherence. Moreover, SCED methodology enabled detailed examination of individual outcome patterns, which is particularly valuable for early-phase evaluation of technology-based interventions. Replication of favourable usability and acceptability findings across independent SCEDs further supports the feasibility of this intervention. While generalisability remains limited due to small sample size, aggregating evidence across similar multiple-baseline studies and progressing to well-controlled randomised trials will be essential to establish definitive efficacy.

### Conclusions

This study provides promising evidence that the PainWaive EEG neurofeedback intervention can reduce pain severity and pain interference in some individuals with CLBP. The variability observed across participants highlights the importance of individual differences in response to neuromodulatory interventions. By prioritising accessibility, usability, and self-administration, this work supports a foundation for more patient-centred, technology-enabled approaches to chronic pain management. Further evaluation of this approach in RCTs is required to establish definitive efficacy.

## AUTHOR CONTRIBUTIONS

**NH-S:** Conceptualization, Methodology, Investigation, Analysis, Validation, Visualization, Supervision, Project administration, Funding, Writing - original draft.

**EE:** Methodology, Investigation, Project administration, Analysis, Writing - review and editing.

**NC:** Investigation, Validation, Writing - review and editing.

**PZ:** Methodology, Investigation, Project administration, Writing - review and editing.

**KC:** Methodology, Software, Writing - review and editing.

**C-TL:** Conceptualization, Funding, Writing - review and editing.

**TN-J:** Conceptualization, Funding, Writing - review and editing.

**SMG:** Conceptualization, Methodology, Investigation, Funding, Supervision, Writing - review and editing.

## ACKNOWLEDGEMENT

The authors acknowledge Sebastian Restrepo, Angus McIntyre, and Lara Cashin for their great contribution to the design and development of the PainWaive Gen-2 EEG headset. We are grateful to all participants who generously contributed their time and insights to this study. We also thank members of the broader research team who provided feedback during the development of the intervention.

## DATA AVAILABILITY

De-identified data will be made available upon reasonable request to the corresponding author.

